# Trends in Hallucinogen-associated Emergency Department Visits and Hospitalizations in California from 2016–2021

**DOI:** 10.1101/2023.08.18.23294275

**Authors:** Steven Tate, Nicolas Garel, Kristin Nash, Anna Lembke

## Abstract

**Objectives:** Hallucinogens encompass a diverse range of compounds with increasing scientific and public interest. Risks associated with hallucinogen use are poorly understood. This study aims to evaluate the trends in hallucinogen-related emergency department visits and hospitalizations in California from 2016 to 2021, as compared to alcohol and cannabis visits and hospitalizations

**Methods:** We conducted a descriptive study on publicly available data on diagnosis codes associated with emergency department (ED) visits and hospitalizations from the California Department of Healthcare Access and Information (HCAI) to describe hallucinogen, alcohol, and cannabis associated encounters.

**Results:** Rates of hallucinogen associated ED visits increased by 69% between 2016 and 2021 compared to a 24% decrease and 1.9% decrease in rates of alcohol associated and cannabis associated ED visits respectively. Rates of hallucinogen associated hospitalizations increased by 74% compared to a 11% increase in alcohol associated hospitalizations and a 19% increase in cannabis associated hospitalizations.

**Conclusions:** The large relative, but small absolute, increase in hallucinogen associated ED visits and hospitalizations is concerning given changing public perception and increasing use of these substances. The categorization of a large number of compounds as hallucinogens limits our ability to understand trends in harms with ICD-10 codes. Improved systems for surveillance of potential harms of hallucinogen use are needed.

## Introduction

Hallucinogens, as defined by the *Diagnostic and Statistical Manual of Mental Disorders, Fifth Edition (DSM-5)*^1^, and *International Classification of Diseases, 10th Revision* (ICD-10)^2^, represent a diverse group of compounds including psychedelics (e.g. psilocybin, lysergic acid diethylamide[LSD], mescaline), psychostimulants (e.g. 3,4-Methylenedioxymethamphetamine [MDMA or ecstasy]), and dissociative anesthetics (e.g. phencyclidine [PCP], ketamine). Over the last several decades, hallucinogens have generated important scientific and public interest, with a growing number of published studies on their potential therapeutic use for depression, post-traumatic stress disorder, and other psychiatric disorders^3^. Many states are taking steps to loosen restrictions around personal and therapeutic hallucinogen use, particularly psychedelics. Oregon and Colorado recently passed and implemented ballot measures that allow for use of psilocybin and in California, Senate Bill 58, proposes loosening restrictions around mescaline, ibogaine, Dimethyltryptamine (DMT), and psilocybin^4^.

As studies of reported benefit multiply, the reported risks associated with hallucinogen use vary greatly. There is a growing perception that hallucinogens are safe and adverse events rare^5^. Data from the Monitoring the Future study shows that non-LSD hallucinogen use among young adults aged 18-30 doubled between 2018 and 2021, with 8% of participants reporting past-year use of Hallucinogens in 2021^6^. This represents the highest prevalence of hallucinogen use among young adults since the survey began 35 years ago and follows three decades of stable prevalence. Despite growing interest in their recreational, spiritual, and therapeutic use, our understanding of the harms of hallucinogens, and particularly classical psychedelics, is lacking. The aims of this descriptive study are to evaluate the trends in hallucinogen-associated ED visits and hospitalizations in California between 2016 and 2021, as compared to the same trends for two commonly used substances, alcohol and cannabis.

## Methods

We conducted a descriptive study using the California Department of Healthcare Access and Information (HCAI) data over a specified period of 6 years (2016-2021). The HCAI makes publicly available the diagnosis codes associated with emergency department (ED) visits and hospital discharges. Available data consist of counts of the ICD-10 codes used as either a primary diagnosis code or secondary diagnosis code recorded for billing for an ED visit or hospital discharge. HCAI collects these data as part of their routine monitoring for all healthcare facilities licensed in California. No individual patient level data, or data aggregated by age, sex, or other demographic factors is publically available from HCAI. This study used HCAI “Patient Discharge Data” and “Emergency Department Data” for years 2016 to 2021 published on their data access portal^7^. The ED data include all patients who were seen in an ED in California and then discharged and does not include patients who were admitted to the hospital from the ED.

2016 was chosen as the index year given the change in diagnosis code use from ICD-9 to ICD-10 in October of 2015. Data were analyzed using Stata 17. Diagnosis codes were classified as hallucinogen, cannabis, or alcohol associated if they were from the corresponding block of “mental and behavioral disorders due to psychoactive substance use” (F16, F10, F12 for hallucinogens, cannabis, and alcohol respectively). Each instance of the code represented one ED visit or hospitalization given CMS coding guidelines that only allow one use disorder associated code per substance for an encounter. Poisoning or overdose codes associated with hallucinogens, cannabis, or alcohol (from the T36-T65 blocks) were not included in the counts since there was potential for an individual visit to have both a use disorder code and overdose code for one visit.

Rates per 10,000 ED visits or hospitalizations were calculated based on the number of total ED visits or hospitalizations per year. For each rate, the % change from 2016 (the index year) was assessed. This study was reported using the guidelines presented in the REporting of studies Conducted using Observational Routinely-collected Data (RECORD) extension of the STrengthening the Reporting of OBservational studies in Epidemiology (STROBE) Guidelines^8^. IRB approval was not necessary given the data were publicly available.

## Results

Table 1 lists the counts of ED visits and hospitalizations in California that were hallucinogen, cannabis, and alcohol associated. In 2016 there were 2,260 ED visits and 2,556 hospitalizations that were hallucinogen associated compared to 4,161 ED visits and 4,126 hospitalizations that were hallucinogen associated in 2021 (Figure 1A), representing an 84% and 61% increase respectively. When accounting for change in total ED visits, the rate of hallucinogen associated visits increased from 2.3 ED visits per 10,000 ED visits in 2016 to 3.8 visits per 10,000 ED visits in 2021, a 69% increase. Similarly the rate of hallucinogen associated hospitalizations increased from 6.7 per 10,000 hospitalizations to 11.6 per 10,000 hospitalizations, a 74% increase (Figure 1B). By contrast, the rate of alcohol-associated ED visits was 328.8 per 10,000 ED in 2016 compared to 256.5 per 10,000 in 2021, representing an 11% increase. For cannabis, the rates of cannabis associated ED visits in 2016 was similar to 2021 (121 visits per 10,000 ED visits to 118.6 visits per 10,000 ED visits), whereas the cannabis associated hospitalization rate increased 19% during the same time period from 288 hospitalizations per 10,000 hospitalizations to 342.3 hospitalizations per 10,000 hospitalizations. Figure 1C compares the percent change in rates for each substance.

**Table 1.** Counts, rates per 10,000, and % change from 2016 of Hallucinogen-associated, Alcohol-associated, and Cannabis-associated Emergency Department Visits and Hospitalizations from 2016-2021.

| Year | <u>Hallucinogen-associated</u> |  |  | <u>Alcohol-associated</u> |  |  | <u>Cannabis-associated</u> |  |  |
| --- | --- | --- | --- | --- | --- | --- | --- | --- | --- |
|  | Count | rate per<br>10,000<br>visits | % change<br>from 2016 | Count | rates per<br>10,000<br>visits | % change<br>from 2016 | Count | rates per<br>10,000<br>visits | % change<br>from 2016 |
| 2016 | 2,260 | 2.3 | 0.0 | 326,550 | 325.8 | 0.0 | 121,267 | 121.0 | 0.0 |
| 2017 | 2,697 | 2.1 | -7.1 | 298,083 | 231.4 | -29.0 | 133,002 | 103.2 | -14.7 |
| 2018 | 2,940 | 2.3 | 3.0 | 294,319 | 232.5 | -28.6 | 136,416 | 107.7 | -10.9 |
| 2019 | 3,419 | 2.7 | 18.0 | 296,017 | 230.4 | -29.3 | 143,027 | 111.3 | -8.0 |
| 2020 | 4,196 | 4.2 | 85.7 | 265,997 | 265.4 | -18.5 | 131,727 | 131.4 | 8.6 |
| 2021 | 4,161 | 3.8 | 69.2 | 268,854 | 246.5 | -24.3 | 129,404 | 118.6 | -1.9 |

| Year | <u>Hallucinogen-associated</u> |  |  | <u>Alcohol-associated</u> |  |  | <u>Cannabis-associated</u> |  |  |
| --- | --- | --- | --- | --- | --- | --- | --- | --- | --- |
|  | Count | rate per<br>10,000<br>admits | % change<br>from 2016 | Count | rate per<br>10,000<br>admits | % change<br>from 2016 | Count | rate per<br>10,000<br>admits | % change<br>from 2016 |
| 2016 | 2,556 | 6.7 | 0.0 | 224,064 | 583.1 | 0.0 | 110,683 | 288.0 | 0.0 |
| 2017 | 2,700 | 7.0 | 5.3 | 221,648 | 574.8 | -1.4 | 124,203 | 322.1 | 11.8 |
| 2018 | 2,898 | 7.6 | 14.1 | 228,246 | 597.6 | 2.5 | 128,455 | 336.3 | 16.8 |
| 2019 | 3,038 | 7.9 | 19.3 | 231,968 | 606.1 | 3.9 | 122,747 | 320.7 | 11.3 |
| 2020 | 3,986 | 11.6 | 74.0 | 222,943 | 647.2 | 11.0 | 117,604 | 341.4 | 18.5 |
| 2021 | 4,126 | 11.6 | 74.4 | 229,550 | 645.2 | 10.7 | 121,769 | 342.3 | 18.8 |

**Figure 1.**
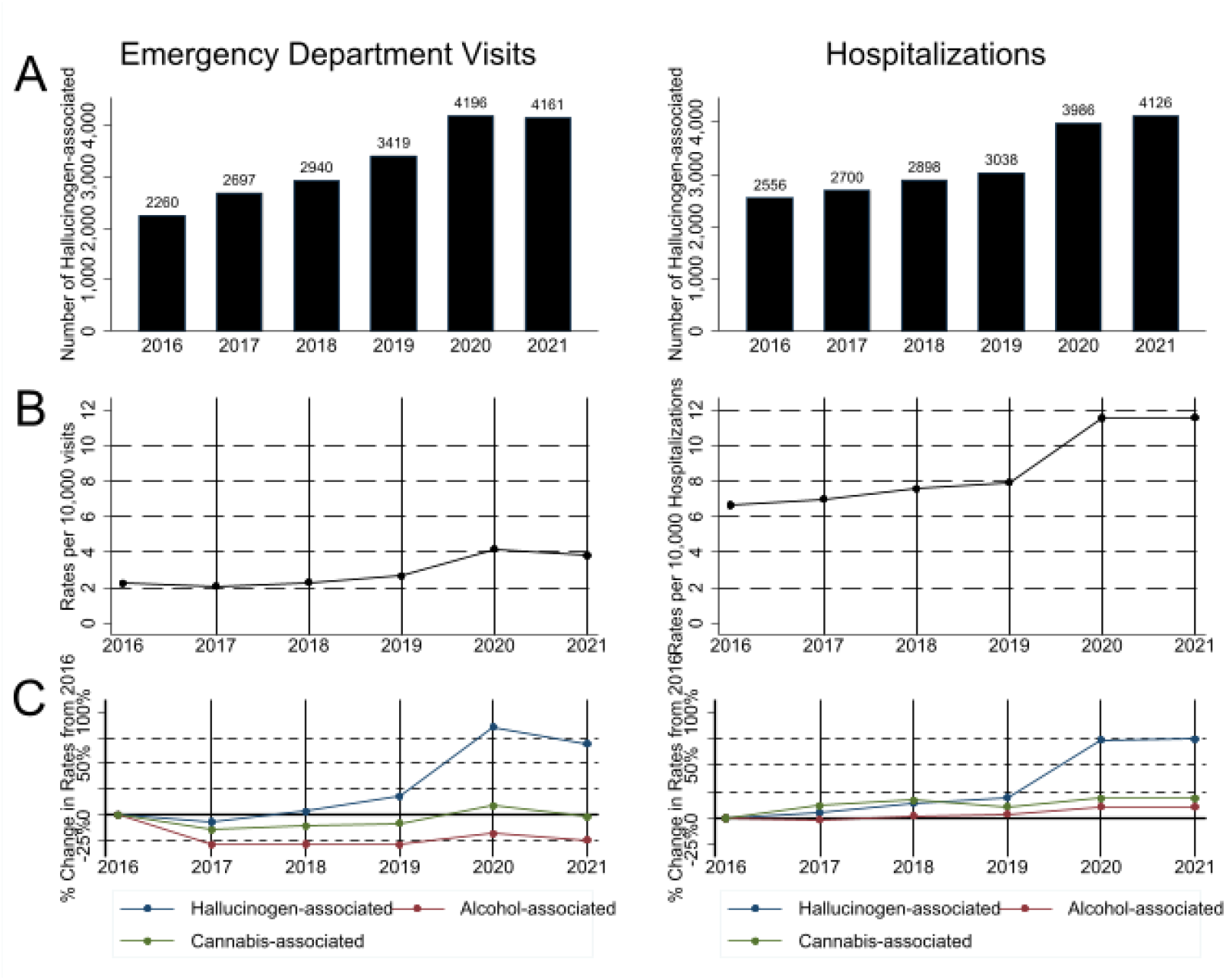
(A) Count of emergency department visits associated with hallucinogen use, (B) rate per 10,000 emergency department visits, (C) % change since 2016 of hallucinogen associated ED visits (blue), alcohol associated ED visits (red), and cannabis associated (green).

## Discussion

Hallucinogen-related ED visits and hospitalizations increased 69% and 74% respectively in California from 2016 to 2021. Although a small absolute increase, these findings represent a concerning trend, especially in light of growing public perception that hallucinogens are safe. This important relative increase was not observed in alcohol and cannabis related visits.

Current data on hallucinogen risks stem primarily from short-term, controlled clinical trials where participants are screened out for psychiatric comorbidities and other risk factors and hence not necessarily representative of the types of individuals who are most likely to use hallucinogens outside of a clinical setting. Further, a substantial dearth of comprehensive public health data concerning hallucinogens makes it difficult to assess their impact at the population level.

Hallucinogen use, as categorized within the ICD-10 codes, encompasses a spectrum ranging from psychedelics to MDMA to ketamine. However, these substances diverge significantly in terms of pharmacology, tolerability, and safety profiles^9^. Aggregating these diverse drugs into one category makes it difficult to assess the changing landscape of use and related harms. The absence of standardized and comprehensive reporting systems to track the harms associated with these substances further compounds the challenge. As such, there is a pressing need for broader and more rigorous tracking of the harms associated with these emerging substances on the individual and population level, especially as policy changes increase access.

### Limitations

A significant limitation of this study is that the data publicly available represents only counts of ICD-10 diagnosis codes associated with ED visits and hospitalization. The lack of additional information on these visits and hospitalizations, such as demographics, precludes a more detailed analysis of these trends observed. The ICD-10 codes are generated for billing purposes and not primarily for research or public health surveillance. These codes depend on doctors recognizing hallucinogen associated disorders.

## Conclusion

Hallucinogen-associated emergency visits and hospitalizations increased between 2016-2021, though the absolute number of visits and hospitalizations remains small compared to alcohol and cannabis. More research on hallucinogen related harms is needed to inform public health and policy discussions about personal and therapeutic use of these compounds.

## Data Availability

All data produced are available online at https://hcai.ca.gov/data-and-reports/datasets

## Acknowledgements

We thank Leonard Nash who performed the preliminary analysis of this data that informed this work.

## Supplemental Digital Content

None

## References

1. Diagnostic and Statistical Manual of Mental Disorders. DSM Library. Accessed August 18, 2023. https://dsm.psychiatryonline.org/doi/book/10.1176/appi.books.9780890425787

2. ICD-10 Version:2019. Accessed August 18, 2023. https://icd.who.int/browse10/2019/en

3. Yaden DB, Potash JB, Griffiths RR. Preparing for the Bursting of the Psychedelic Hype Bubble. JAMA Psychiatry. 2022;79(10):943–944. doi:10.1001/jamapsychiatry.2022.2546

4. Wiener S, Kalra A, Becker J, et al. SB-58 Controlled substances: decriminalization of certain hallucinogenic substances. California Legislative Information. Accessed August 10, 2023. https://leginfo.legislature.ca.gov/faces/billTextClient.xhtml?bill_id=202320240SB58

5. Livne O, Shmulewitz D, Walsh C, Hasin DS. Adolescent and adult time trends in US hallucinogen use, 2002–19: any use, and use of ecstasy, LSD and PCP. Addict Abingdon Engl. 2022;117(12):3099–3109. doi:10.1111/add.15987

6. Keyes KM, Patrick ME. Hallucinogen use among young adults ages 19–30 in the United States: Changes from 2018 to 2021. Addiction. n/a(n/a). doi:10.1111/add.16259

7. Data and Reports. California Department of Healthcare Access and Information. Accessed August 16, 2023. https://hcai.ca.gov/data-and-reports/

8. Benchimol EI, Smeeth L, Guttmann A, et al. The REporting of studies Conducted using Observational Routinely-collected health Data (RECORD) Statement. PLOS Med. 2015;12(10):e1001885. doi:10.1371/journal.pmed.1001885

9. Gregorio DD, Aguilar-Valles A, Preller KH, et al. Hallucinogens in Mental Health: Preclinical and Clinical Studies on LSD, Psilocybin, MDMA, and Ketamine. J Neurosci. 2021;41(5):891–900. doi:10.1523/JNEUROSCI.1659-20.2020

